# Epidemiology, clinical presentation and management of COVID-19 associated Mucormycosis: A single center experience from Pune, Western India

**DOI:** 10.1101/2021.09.15.21263622

**Authors:** Ameet Dravid, Reema Kashiva, Zafer Khan, Balasaheb Bande, Danish Memon, Aparna Kodre, Prashant Potdar, Milind Mane, Asir Tamboli, Vishal Pawar, Dattatraya Patil, Debashis Banerjee, Kailas Bhoite, Akshay Shinde, Reshma Pharande, Suraj Kalyani, Prathamesh Raut, Madhura Bapte, Charlotte Saldanha, Dinesh Chandak, Fouzia Ajani, Anshul Mehta, M Sateesh Reddy, Krushnadas Bhayani, S S Laxmi, P D Vishnu, Shipra Srivastava, Shubham Khandelwal, Sailee More, Atif Shakeel, Mohit Pawar, Pranava Nande, Amol Harshe, Sagar Kadam, Sudhir Hallikar, Nudrat Kamal, Danish Andrabi, Sachin Bodhale, Akshay Raut, Pushkar Gawande, Ankush Bhandari, Nilesh Wasmatkar, Adnanali Sarkar, Sangeeta Chandrashekhar, Pallavi Butiyani, Geetanjali Akhade, Aditi Abnave, Chandrashekhar Raman, Siraj Basade, Uma Mahajan, Gaurav Joshi, Dilip Mane

## Abstract

**Background:** The second COVID-19 wave in India, triggered by the Delta variant,has been associated with an unprecedented increase in cases of COVID-19 associated Mucormycosis(CAM), mainly Rhino-orbito-cerebral mucormycosis(ROCM).The primary reason appears to be an unusual alignment of multiple risk factors in patients like prevalence of hypoxia, uncontrolled diabetes mellitus, indiscriminate use of steroids, high iron levels and immune dysfunction.

**Methods:** This retrospective cohort study was conducted at Noble hospital and Research Centre (NHRC), Pune, Western India between 1^st^ April 2020 and 1^st^ August 2021 to identify patients admitted with CAM. The primary endpoint was incidence of all cause mortality due to CAM. Secondary outcomes studied were need for mechanical ventilation and intensive care unit(ICU) admission. Baseline and time dependent risk factors significantly associated with death due to CAM were identified by Relative risk estimation.

**Results:** 59 patients were diagnosed with Mucormycosis at NHRC (58 ROCM, 1 Renal (disseminated) mucormycosis). Median age of the cohort was 52(IQR: 41,61) years and it included 20.3% females. Median duration from first positive COVID-19 RT PCR test to diagnosis of Mucormycosis was 17(IQR: 12,22) days. 90% patients were diabetic with 30% being newly diagnosed at the time of COVID-19 admission and 89% having uncontrolled sugar level (HbA1c > 7%). All patients were prescribed steroids during treatment for COVID-19. 56% patients were prescribed steroids for non-hypoxemic, mild COVID (irrational steroid therapy) while in 9%, steroids were indicated but were prescribed in inappropriately high dose. Majority of the patients were treated with a combination of surgical debridement(94%), intravenous Amphotericin B(91%) and concomitant oral Posoconazole therapy(95.4%). 74.6% patients were discharged after clinical and radiologic recovery while 25.4% (15 patients) died. On Relative risk analysis, CT severity score during COVID-19 admission ≥18 (p=0.017), presence of orbital symptoms(p=0.002), presence of diabetic ketoacidosis(p=0.011) and cerebral involvement by Mucor(p=0.0004) were associated with increased risk of death. Duration of Amphotericin B therapy of ≥ 21 days was associated with statistically significant reduction in mortality(p=0.002).

**Conclusions:** CAM is an uncommon, rapidly progressive, angioinvasive, opportunistic fungal infection which is fatal if left untreated. Combination of surgical debridement and antifungal therapy leads to clinical and radiologic improvement in majority of cases.

## Introduction

In December 2019, Wuhan city, the capital of Hubei province in China, became the centre of an outbreak of viral pneumonia. By Jan 7, 2020, scientists had isolated a novel RNA, beta coronavirus from these patients. It was named severe acute respiratory syndrome coronavirus 2 (SARS-CoV-2) due to its sequence homology with SARS-COV-1 ^[1]^. The disease caused by SARS-COV-2 was later designated coronavirus disease 2019 (COVID-19) in February 2020, by World health organization (WHO) ^[2]^. COVID-19 spread rapidly worldwide and India was no exception. By 1^st^ August 2021, there have been more than 300 million infections and 0.4 million deaths due to COVID-19 in India ^[3]^. With COVID-19, the incidence of secondary bacterial or fungal infections is 8%, with aspergillosis and candida being the most common fungi reported ^[4,5]^. The second COVID wave in India (March 2021 – May 2021), triggered by the Delta variant, has also been associated with an unprecedented increase in the cases of COVID-19 associated Mucormycosis (CAM) ^[6,7]^. Globally, the prevalence of mucormycosis varies from 0.005 to 1.7 per million population, while its prevalence is nearly 80 times higher (0.14 per 1000) in India as per a recent estimate of year 2019-2020 ^[7-9]^. Thus, India already had the highest cases of mucormycosis in the world and the incidence increased exponentially during the second COVID wave. As of August 1^st^, 2021, 45,374 cases of CAM have been reported in India ^[6]^.

CAM is an uncommon, rapidly progressive, angio-invasive, commonly fatal, opportunistic fungal infection ^[9]^. Unusual alignment of multiple risk factors could be associated with sudden spurt of CAM in India ^[10]^. Dysregulated immune response in COVID-19 characterized by exuberant activation of innate immune system, elevation in systemic inflammatory markers (C-reactive protein (CRP), ferritin, lactate dehydrogenase (LDH) and D-dimer), aberrant pro-inflammatory cytokine secretion (Interleukin – 6 (IL-6), soluble IL-2 receptor [IL-2R], IL-10, TNF-α) by alveolar macrophages and depleted adaptive immune response (decline in CD4+ T cell, CD8+ T cell, Natural killer cell and decreased Interferon Gamma (IFN-γ) expression in CD4+ T cells) could be one of the factors predisposing individuals to CAM ^[11,12]^. In these patients, the activation of antiviral immunity to SARS-CoV-2 may, paradoxically, potentiate an inflammatory phenotype and thus may favor secondary infections. In addition, India has the second largest population with diabetes mellitus (DM) in the world (77 million people with diabetes, with a nationwide prevalence of 7.3 %.)^[13, 14]^. SARS-CoV-2 has been shown to affect the beta cells of the pancreas, resulting in metabolic derangement, possibly causing diabetes mellitus ^[15]^. Uncontrolled DM is a risk factor for both, COVID-19 and Mucormycosis. Steroids, namely Dexamethasone and Methylprednisolone have been extensively used to resolve hyperinflammation and inflammatory lung damage in severe COVID-19 ^[16-18]^. But use of steroids can cause hyperglycemia, suppression of several polymorphonuclear (PMN) leucocyte functions, impairment of phagocytosis by resident macrophages, depletion of T cell immunity ^[10]^ and increase risk of Mucormycosis. Indiscriminate use of steroids in patients with mild or asymptomatic COVID-19 can be counter-productive and make the person susceptible to opportunistic fungi ^[19]^. Use of immunomodulator therapy like Tocilizumab (TCZ, IL-6 receptor inhibitor) in combination with steroids for Cytokine release syndrome (CRS, cytokine storm) could further aggravate immunosuppression and facilitate breakthrough fungal infection ^[20]^. Thus, the primary reason for this sudden increase in CAM in India appears to be germination of Mucorales spores in an ideal environment of low oxygen (hypoxia), high glucose (diabetes mellitus, new onset hyperglycemia, impaired fasting glucose or steroid-induced hyperglycemia), acidic environment^[10]^ (metabolic acidosis, diabetic ketoacidosis [DKA]), high iron levels (hyper-ferritinemia characteristic of COVID-19 leading to excess intracellular iron that generates reactive oxygen species and resultant death of cells with acidosis facilitating free iron release for mucor germination) ^[21,22]^ and immune dysfunction (mediated by Delta variant of SARS COV-2, steroid therapy or background co-morbidities).

The most common form of Mucormycosis seen in India during the COVID-19 pandemic was the Rhino-orbito-cerebral (ROCM) one but cases of pulmonary ^[7,23]^ or disseminated mucormycosis ^[24]^ have also been reported. Suspected ROCM requires urgent intervention, because of the often rapidly progressive and destructive nature of the infection. Delayed initiation of therapy is associated with increased mortality ^[25,26]^. Despite treatment, case-fatality rates due to mucormycosis during the pre-COVID-19 pandemic era were already high, ranging from 32% to 70%, according to organ involvement ^[25,26]^. However, in SARS-CoV-2 infection, the mortality maybe even higher. Maximizing survival rates requires rapid diagnostic and therapeutic intervention, including immediate involvement of a multidisciplinary medical, surgical, radiological, and laboratory-based team. Multiple case reports ^[27-50]^ and retrospective cohort studies ^[7, 51-61]^ of CAM from India and other countries have already been published. However, detailed epidemiology, risk factor analysis, surgical and medical treatment administered, outcomes and complications (especially long-term complications) developing in these patients has been scarcely reported ^[7,52,57]^. In addition, lack of imaging modalities in rural areas of India, inability to identify early symptoms by general practitioners, shortage of antifungal drugs (especially Liposomal Amphotericin B) during second COVID wave and inadequate number of specialists to perform complex endoscopic, orbital and skull base surgeries ^[7,10]^ posed a serious challenge in tackling rising ROCM cases in India and led to a high mortality in certain cohort studies ^[7,52]^. As a result, we planned this retrospective observational cohort study aimed at understanding the epidemiology, clinical presentation, outcomes and long-term complications in patients with CAM admitted at our tertiary level hospital in Pune, Western India during the COVID-19 pandemic.

## Methods

### Study Setting

This retrospective cohort study was conducted at Noble hospital and Research Centre (NHRC), Pune, Western India. NHRC is a tertiary level private hospital designated for clinical management of COVID-19 patients and management of post COVID complications since 23^rd^ March 2020. Pune is located in the state of Maharashtra, Western India and was one of the epicenters of COVID-19 epidemic in India. As of 1^st^ August 2021, Maharashtra state has reported more than 6.3 million cases of COVID-19 and more than 133,000 deaths ^[3]^. Till 1^st^ August 2021, NHRC has admitted 5439 COVID-19 patients with 391 deaths. Maharashtra state has also reported more than 5000 cases of COVID-19 associated Mucormycosis (CAM) till 1^st^ August 2021^[6]^.

NHRC provides clinical care, diagnostic and treatment services to patients at a subsidized cost. Data of all hospitalized patients is entered into an electronic database (Lifeline electronic database, Manorama infosystems, Kolhapur, India).

### Study Population

Patients were eligible for inclusion in this analysis if they were admitted to NHRC between 1^st^ April 2020 and 1^st^ August 2021 and were diagnosed with COVID-19 associated Mucormycosis (CAM). COVID-19 diagnosis was made in patients who tested positive for SARS COV 2 RNA in respiratory specimens by reverse transcription PCR (RT-PCR) or a positive rapid antigen test. Mucormycosis was identified in patients having compatible clinical and radiologic manifestations and demonstration of fungi in the tissue or sterile body fluids by either direct microscopic visualization of broad ribbon-like aseptate hyphae (potassium hydroxide (KOH) mount or Calcofluor stain), culture isolation of Mucorales (Sabouraud dextrose agar) or Histopathology examination of affected tissue showing Mucor species (Hematoxylin and eosin, periodic acid Schiff, or Gomori methenamine silver stain) ^[9]^. CAM was defined as development of Mucormycosis within 3 months of diagnosis of COVID-19.

Data was obtained from electronic health record of each individual admitted for treatment of CAM in NHRC by manual abstraction. It included hospitalization dates, demographics, co-morbidities, severity of associated COVID-19 illness, treatment received for COVID-19 including steroids and immunomodulator therapy, presenting symptoms of Mucormycosis, clinical examination data, anatomic site of involvement, diagnostic modalities including microscopy, culture, or histopathology, laboratory investigation data (including inflammatory markers), microbiology reports, imaging reports (High resolution computerized tomography scan of chest, paranasal sinuses and brain (HRCT chest, CT PNS and CT brain)), treatment details, including antifungal drug therapy and surgical debridement, data on use of supplemental oxygen, mechanical ventilation and hospitalization outcomes. Mild COVID was defined as individuals who have various signs and symptoms of COVID-19 but who do not have shortness of breath, dyspnea, or abnormal chest imaging ^[62]^. Moderate disease was defined as patients who show evidence of lower respiratory disease during clinical assessment or imaging and who have oxygen saturation (SpO_2_) 90 to ≤ 93% on room air. Severe COVID was defined as SpO_2_ < 90% on room air, a ratio of arterial partial pressure of oxygen to fraction of inspired oxygen (PaO_2_/FiO_2_) < 300 mm Hg, respiratory frequency > 30 breaths/min or lung infiltrates >50% ^[62]^. Critical COVID was defined as presentation with respiratory failure, septic shock, and/or multiple organ dysfunction. For all patients diagnosed with CAM, we also scrutinized inpatient case files until hospital discharge, death, or 1^st^ September 2021—the date on which the database was locked—whichever happened first. Systemic complications developing in patients during hospital admission were also noted. All patients who died due to CAM during hospital admission were identified and a death audit to look for complications and cause of death was undertaken. For patients who left the hospital against medical advice, we considered a worst-case scenario for mortality analysis and assumed the patients died. All patients who recovered, got discharged from NHRC and had outpatient follow-up at 15, 30 and 90 days after discharge were identified. Their outpatient follow-up visits were traced from electronic database to look for delayed complications.

### Management of Mucormycosis at NHRC

NHRC protocol for management of Mucormycosis has been developed after careful consideration of current global and national guidelines ^[63,64]^. As per the protocol, early, radical, surgical debridement of affected site in addition to combination systemic antifungal therapy was utilized for treatment.

### Surgical management

Functional endoscopic paranasal sinus surgery (FESS) with debridement of necrosed and diseased sinus tissue and orbital decompression was the most common surgical procedure performed in patients with ROCM. All efforts were made to preserve the eye for as long as possible in view of role of eye to not only provide vision but also its removal causing significant psycho-social problems to patient. In patients with total blindness, proptosis, fixed pupil and eyeball, imaging evidence of orbital involvement by Mucor (globe/muscles/fat) and/or intracranial spread (superior orbital fissure/inferior orbital fissure involvement), surgical exenteration of the eye was performed ^[65]^. Surgical exenteration of the eye with debridement of orbital cavity was performed in consultation with Department of Otorhinolaryngology and Ophthalmology. Partial or total maxillectomy was performed in patients with evidence of osteomyelitis of maxilla or alveolar arch. Neurosurgery was performed in case of intracranial spread of disease with involvement of skull base or formation of basi-frontal lobe brain abscess.

### Medical management

Combination therapy of Liposomal Amphotericin B (LAmB) and Oral Triazole (predominantly oral Posaconazole) was the most common medical treatment offered to all patients with CAM. LAmB was initiated at the dose of 5 mg/kg. The dose was increased to 10 mg/kg in case of intracranial spread ^[63,64]^. In the scenario of shortage of LAmB due to surge in CAM cases or non affordability of patient, other formulations of Amphotericin B (Amphotericin B lipid complex (5 mg/kg), Amphotericin B lipid emulsion (5 mg/kg) and Amphotericin B de-oxycholate (1 mg/kg)) were used for patients. The duration of Amphotericin B treatment was decided depending on the site of involvement (3 weeks for only paranasal sinus involvement, 4 weeks for orbital, lung and disseminated Mucormycosis and 6 weeks for Central nervous system (CNS) Mucormycosis). Oral Posaconazole (delayed release tablets, 300 mg twice a day on Day 1 and then 300 mg once a day) was started along with Amphotericin B treatment and continued after discharge from hospital for a period of 1 to 3 months depending on local control of disease and discretion of Infectious disease physician. In patients with concomitant chronic kidney disease, intravenous Posaconazole or Isavuconazole followed by step-down to oral therapy was treatment of choice. Serum Posaconazole levels were performed to guide dosage during oral therapy.

### Outcomes

Primary endpoint:

1. Deaths in the cohort due to CAM.

Secondary outcomes:

2. Number of patients who had a clinical and radiologic recovery and were discharged from hospital.
3. Patients who required mechanical ventilation (noninvasive or invasive) and Intensive care unit (ICU) admission for CAM.
4. Incidence of systemic complications (including long-term complications) in patients after starting Amphotericin B therapy. The use of database for clinical research was approved by the institutional review board (IRB) of Noble hospital and Research Centre, Pune, India.

## Statistical Methods

Continuous variables were summarized using median and interquartile range (IQR), while categorical variables were summarized using frequency and percentages. Continuous variables were compared using a Mann Whitney U test. Categorical variables were compared using Chi-square test, Proportion test and Fishers’ exact test. Baseline and time dependent risk factors significantly associated with death due to CAM were identified by Relative risk analysis. Baseline risk factors included for analysis were age (< 60 years or ≥ 60 years), gender, severity of baseline COVID disease (CT severity index ≥ 18 versus < 18), presence of orbital or central nervous system (CNS) symptoms, presence of diabetic keto-acidosis, presence of intracranial spread (cerebral involvement) and baseline investigations like absolute lymphocyte count (< 1000 versus ≥ 1000 cells/mm^3^), C reactive protein (≥ 50 versus < 50 mg/L), D-dimer (≥ 252 versus < 252 mcg/mL) and Ferritin (≥ 1000 versus < 1000 mcg/L). Time dependent risk factors included duration of Amphotericin B therapy (< 3 weeks versus ≥ 3 weeks) and development of systemic complications like acute kidney injury, hepatitis, anemia, thrombocytopenia, osteomyelitis and CNS complications (stroke or cerebritis). The p value ≤ 0.05 was considered as statistically significant. All data was analyzed by SPSS version 12.0.

## Results

During the period between 1^st^ April 2020 to 1^st^ August 2021, 59 patients were diagnosed with Mucormycosis at NHRC. Fifty-eight patients were diagnosed to have ROCM while 1 patient was diagnosed to have Renal (disseminated) mucormycosis. Median age of the cohort was 52 (IQR: 41, 61) years and it included 20.3 % (12/59) females. Seventeen (28.8 %) patients were > 60 years of age. Fifty-six (95 %) patients had pre-existing or newly diagnosed co-morbidities. Diabetes mellitus (53/59, 89.8 %) was the most common co-morbidity seen in our cohort. Of these, 15 patients (28.3 %) were newly diagnosed with diabetes during hospital admission for COVID-19. Two patients were diagnosed to have impaired fasting glucose (Fasting blood glucose between 110 and 125 mg/dl). Preexisting co-morbidities observed in patients at admission are enumerated in Table 1. Three patients (5.1%) had no co-morbidities prior to developing CAM.

**Table 1:** Epidemiology of COVID-19 admission in Mucormycosis patients.

| Baseline characteristics | Total<br>(N=59)<br>n (%) | Discharged<br>(n=44)<br>n (%) | Death<br>(n=15)<br>n (%) | p-value |
| --- | --- | --- | --- | --- |
| **Age (Years)** |  |  |  | 0.077 |
| ≤ 60 years | 42 (71.2) | 34 (77.3) | 8 (53.3) |  |
| > 60 years | 17 (28.8) | 10 (22.7) | 7 (46.7) |  |
| Median (IQR) | 52 (41, 61) | 50 (40, 58) | 57 (42, 65) | 0.136 |
| **Gender** |  |  |  | 0.481 |
| Male | 47 (79.7) | 36 (81.8) | 11 (73.3) |  |
| Female | 12 (20.3) | 8 (18.2) | 4 (26.7) |  |
| **Co-morbidities** |  |  |  |  |
| Diabetes Mellitus |  |  |  | 0.216 |
| Non-diabetic | 6 (10.2) | 5 (11.4) | 1 (6.7) |  |
| Known Diabetes Mellitus | 36 (61.0) | 24 (54.5) | 12 (80.0) |  |
| Newly diagnosed DM/ IFG | 17 (28.8) | 15 (34.1) | 2 (13.3) |  |
| Hypertension | 24 (40.7) | 17 (38.6) | 7 (46.7) | 0.585 |
| Chronic kidney disease | 9 (15.3) | 6 (13.6) | 3 (20.0) | 0.680 |
| Ischemic Heart Disease | 7 (11.9) | 4 (9.1) | 3 (20.0) | 0.355 |
| Chronic liver disease | 2 (3.4) | 0 (0.0) | 2 (13.3) | 0.061 |
| Stroke | 1 (1.7) | 0 (0.0) | 1 (6.7) | 0.254 |
| HIV/ Hepatitis B infection | 1 (1.7) | 1 (2.3) | 0 (0.0) | >0.999 |
| Obesity | 2 (3.4) | 1 (2.3) | 1 (6.7) | 0.447 |
| Hypo/hyperthyroidism | 3 (5.1) | 2 (4.5) | 1 (6.7) | >0.999 |
| **Severity of COVID-19** |  |  |  | 0.223 |
| Mild | 16 (27.1) | 12 (27.3) | 4 (26.7) |  |
| Moderate | 29 (49.2) | 24 (54.5) | 5 (33.3) |  |
| Severe | 14 (23.7) | 8 (18.2) | 6 (40.0) |  |
| **HRCT chest COVID severity score** |  |  |  | **0.047** |
| < 18 | 55 (93.2) | 43 (97.7) | 12 (80.0) |  |
| ≥ 18 | 4 (6.8) | 1 (2.3) | 3 (20.0) |  |
| **Median (IQR)** | **12 (8, 15)** | **12 (8, 14)** | **14 (8, 17)** | **0.152** |
| **COVID-19 admission Median(IQR)** |  |  |  |  |
| Duration of hospitalization (days) | 9 (6, 14) | 8 (6, 13.5) | 10 (7, 15) | 0.149 |
| Duration of ICU stay (days) | 0 (0, 10) | 0 (0, 0) | 0 (0, 10) | 0.303 |
| Duration of ventilation (days) | 0 (0, 10) | 0 (0, 0) | 0 (0, 10) | **0.046** |
| **COVID-19 vaccine status** |  |  |  | 0.826 |
| No vaccination | 46 (78.0) | 34 (77.3) | 12 (80.0) |  |
| AstraZeneca - 1 dose | 9 (15.3) | 7 (15.9) | 2 (13.3) |  |
| AstraZeneca - 2 doses | 2 (3.4) | 1 (2.3) | 1 (6.7) |  |
| Covaxin (Bharat Biotech) - 1 dose | 1 (1.7) | 1 (2.3) | 0 (0.0) |  |
| Covaxin (Bharat Biotech)- 2 dose | 1 (1.7) | 1 (2.3) | 0 (0.0) |  |

DM- Diabetes mellitus; IFG- Impaired fasting glucose; HRCT- High resolution computerized tomography; HIV- Human immunodeficiency virus

Clinically, 16 patients (27.1%) had mild COVID, 29 (49.2 %) had moderate COVID and 14 patients (23.7%) had severe or critical COVID disease prior to diagnosis of Mucormycosis. All 59 patients underwent High resolution CT (HRCT) imaging of chest (GE Optima, 128 slice CT scanner) during admission for COVID-19 and median CT severity index was 12 (IQR: 8,14.5). CT severity index ^[66]^ indicated mild disease (CT severity index: < 8) in 18.6 % (11/59), moderate disease (CT severity index: 8-14) in 56 % (33/59) and severe disease (CT severity index: 15-25) in 25.4 % (15/59) of patients. Treatments given during COVID-19 admission are enumerated in Table 2a and 2b. Remdesivir was prescribed to 67.8 % (40/59), intravenous and/or oral steroids were prescribed to 100 % and Immune-modulator therapy (Tocilizumab - 5, Barcitinib-2, Idofinib-1, Infliximab-2, Bevacizumab - 2 and Itolizumab - 1) was prescribed to 22 % (13/59) patients in our cohort. Methylprednisolone was the commonest prescribed steroid (54 patients, 91.5%, Table 2b). Most commonly prescribed dose of Methylprednisolone was 1-2 mg/kg/day in divided doses for 5 to 10 days ^[67]^. Ten patients (16.9 %) were prescribed 2 different steroids during treatment course for COVID-19. 56 % (33/59) patients were prescribed steroids in the absence of hypoxia (irrational or unscientific steroid use). In 5 (5/54, 9.3%) patients, Methylprednisolone dose of more than 2 mg/kg/day was prescribed for treatment of severe COVID-19 (wrong or excess steroid dose). Eleven patients (18.6%) required ICU admission while 7 (11.9 %) needed noninvasive or invasive ventilation during hospital admission of COVID-19 infection. Thirteen patients (22%) had received COVID vaccination prior to admission (3 patients received 2 vaccine doses and 10 patients received single vaccine dose, Table 1). Five patients (8.5 %) were treated for COVID-19 while in home quarantine and did not require hospital admission. All 5 patients in home quarantine received steroids (irrational or unscientific steroid use) as a part for of their treatment regimen. Median duration of hospital admission for COVID-19 was 9 (IQR: 6, 14) days.

**Table 2a:**
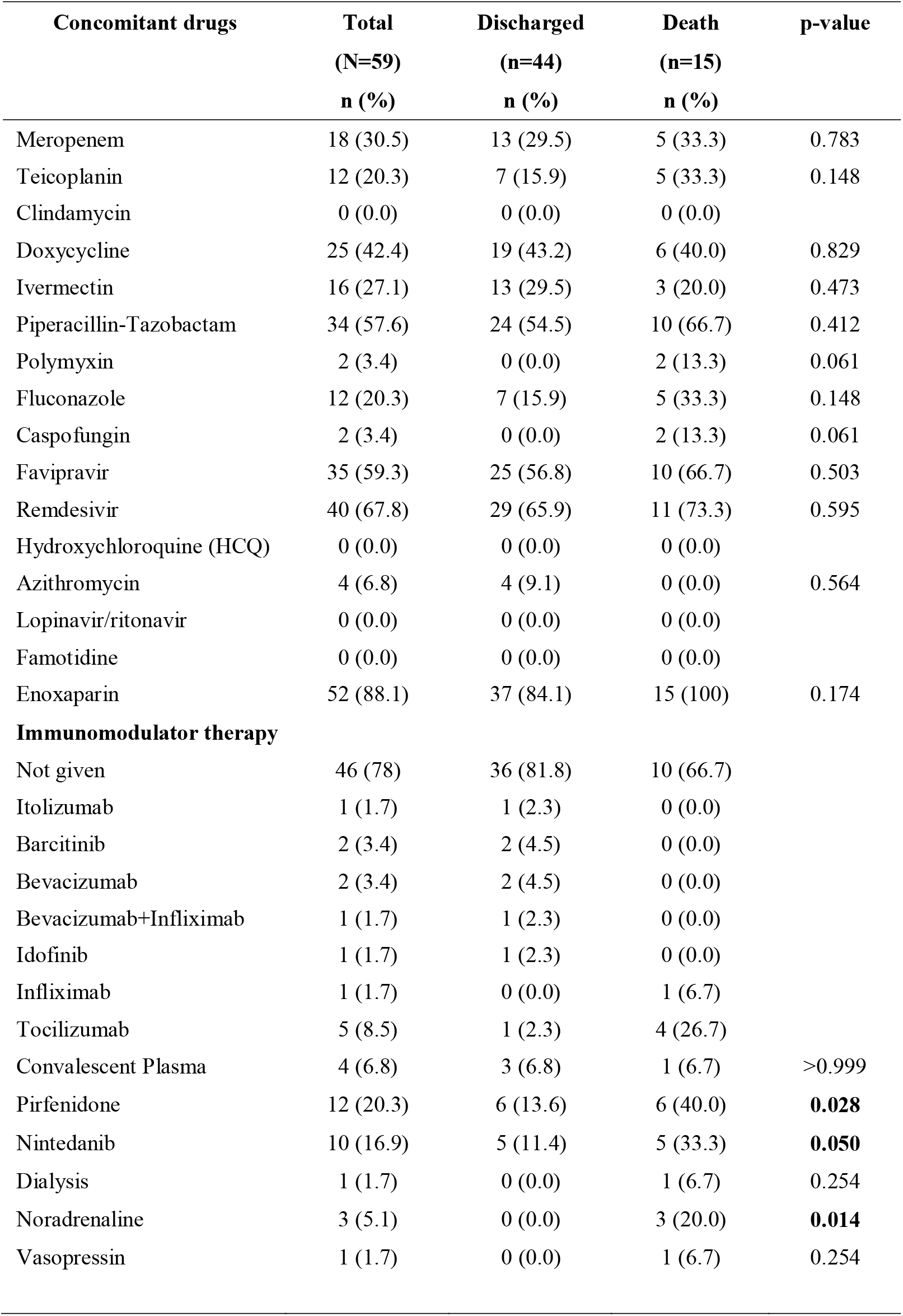
Drugs administered during COVID-19 treatment.

**Table 2b:**
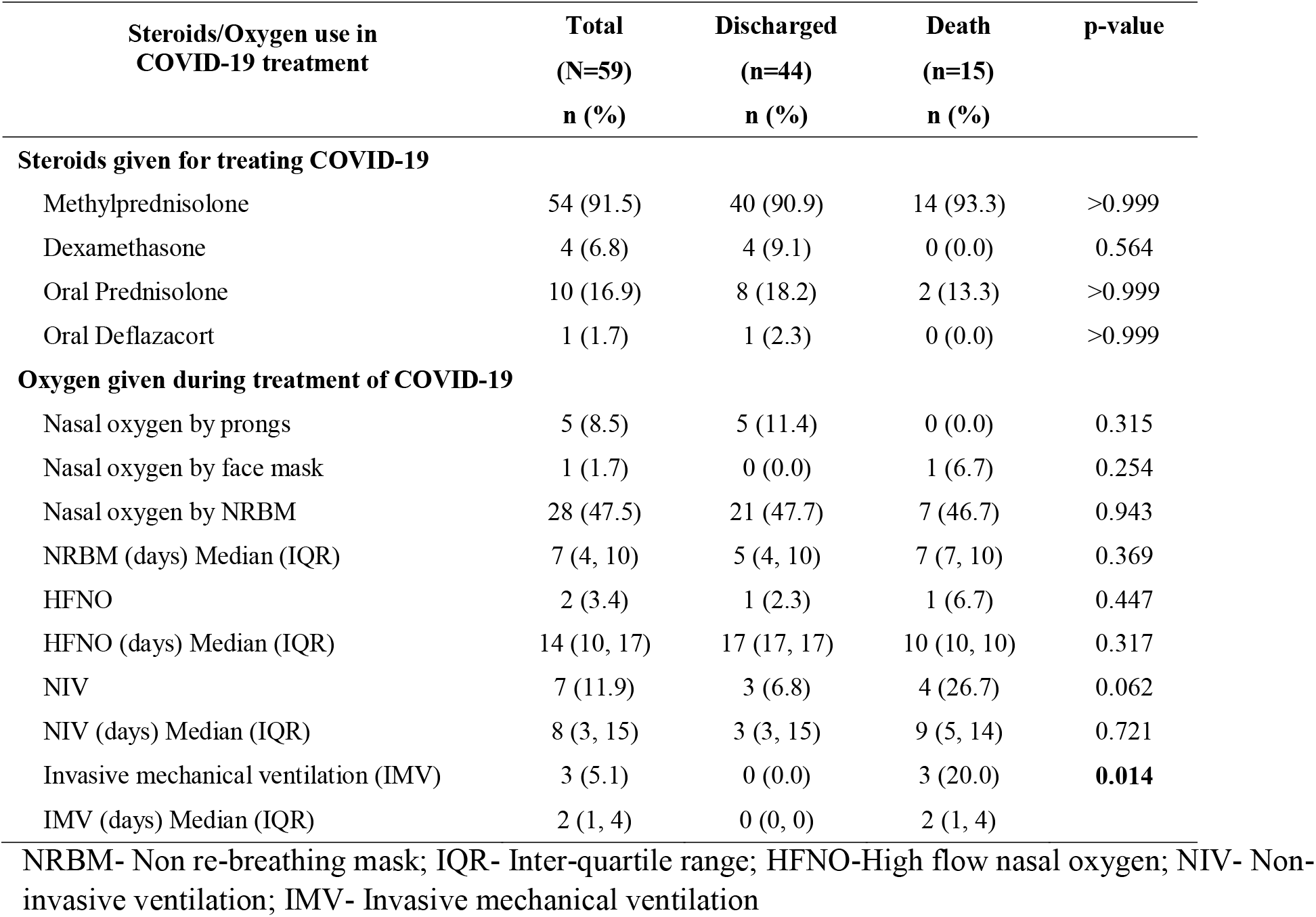
Steroids and Supplemental Oxygen used during COVID-19 treatment.

Median duration from first positive COVID-19 RT PCR test to diagnosis of Mucormycosis was 17 (IQR: 12, 22) days. Duration was less than 14 days for 22 (37.3 %, early CAM) patients while it was > 14 days for 37 (62.7 %, late CAM) patients. Median duration between hospital discharge for COVID-19 to re-admission for Mucormycosis was 7 (IQR: 2.5, 15.5) days. Twenty patients (33.9 %) were diagnosed to have Mucormycosis (suspected fungal sinusitis, ROCM) during admission for COVID-19. Twenty-seven (45.7 %) were re-admitted less than 14 days after hospital discharge for COVID-19 while 12 (20.4 %) were re-admitted after more than 14 days. Headache (43/58, 74.1 %), hemi-facial pain predominantly maxillary pain (42/58, 72.4 %) and facial swelling (33/58, 56.9 %) were the three commonest presenting symptoms of ROCM in our cohort (Table 3). Median duration of symptoms was 6 (IQR: 3, 8.5) days. Seven patients (11.9 %) had symptoms for more than 2 weeks prior to diagnosis of ROCM. Fifty-eight patients (99%) had rhinal (paranasal sinus) involvement, 33 (56%) had orbital (eyeball, extra-ocular muscles or orbital cavity) and 26 (44.1%) had cerebral involvement (infarcts, cavernous sinus involvement, dural enhancement, frontal lobe abscess, osteomyelitis and erosion of frontal bone, Table 3). Palatal involvement (palatal necrosis, black eschar or palatal osteomyelitis) was observed in 13 (22 %) patients. Median hemoglobin value at diagnosis of Mucormycosis was 13.4 (IQR: 11.4, 14.5) mg/dl (Table 4). Median absolute neutrophil count (ANC) was 8100 (IQR: 5248, 11475) cells/mm^3^ and median absolute lymphocyte count (ALC) was 1350 (IQR: 1010, 1750) cells/mm^3^. Fourteen patients (24.6 %) had ALC < 1000 cells/mm^3^ while no patient had ANC < 1000 cells/mm^3^ at admission. Median CRP at admission was 84.7 (IQR: 41,167, 45 patients) mg/L, median D-dimer was 483.5 (IQR: 253.7,1201, 37 patients) mcg/mL, and median Ferritin value was 857.1 (IQR: 280.5, 1429, 32 patients) mcg/L. Thirteen patients (40.6 %) had serum Ferritin level > 1000 mcg/L at admission while 11 patients had D-dimer > 1000 mcg/mL. Median HbA1C level in our cohort was 9.3 (IQR: 8, 12). Ten patients (10/53, 18.9%) presented with diabetic ketoacidosis. Diagnosis of Mucormycosis was made by KOH/calcofluor white staining of nasal scrapings (25/59, 42.4%), culture (5/59, 8.5%) and histopathologic examination of excised tissue from paranasal sinuses (57/59, 96.6%). Mixed mould infection (mucormycosis and aspergillosis) was observed in 3 patients.

**Table 3:** Baseline characteristics of patients of Mucormycosis.

| **Mucormycosis** | **Total**<br>**n (%)** | **Discharged**<br>**n (%)** | **Death**<br>**n (%)** | **p-value** |
| --- | --- | --- | --- | --- |
| **Total patients** | **59** | **44 (74.6)** | **15 (25.4)** | **0.0002** |
| **Type of Mucormycosis** |  |  |  | **<0.0001** |
| Rhinal involvement only | 24 (40.7) | 24 (54.5) | 0 (0.0) |  |
| Rhinal + Orbital involvement | 8 (13.6) | 7 (15.9) | 1 (6.7) |  |
| Rhinal + Orbital + Cerebral | 25 (42.4) | 13 (29.5) | 12 (80.0) |  |
| Rhinal + Cerebral | 1 (1.7) | 0 (0.0) | 1 (6.7) |  |
| Renal | 1 (1.7) | 0 (0.0) | 1 (6.7) |  |
| **Symptoms of Mucormycosis** |  |  |  |  |
| Facial pain | 42 (71.2) | 32 (72.7) | 10 (66.7) | 0.654 |
| Facial swelling | 33 (55.9) | 22 (50.0) | 11 (73.3) | 0.142 |
| Headache | 43 (72.9) | 32 (72.7) | 11 (73.3) | >0.999 |
| Eye swelling | 33 (55.9) | 20 (45.5) | 13 (86.7) | **0.007** |
| Eye pain | 29 (49.2) | 17 (38.6) | 12 (80.0) | **0.007** |
| Loss of vision | 21 (35.6) | 9 (20.5) | 12 (80.0) | **<0.0001** |
| Ptosis | 21 (35.6) | 11 (25.0) | 10 (66.7) | **0.004** |
| Deviation of angle of mouth | 5 (8.5) | 3 (6.8) | 2 (13.3) | 0.593 |
| Nasal discharge | 15 (25.4) | 13 (29.5) | 2 (13.3) | 0.310 |
| Limb weakness | 5 (8.5) | 1 (2.3) | 4 (26.7) | **0.013** |
| Toothache | 27 (45.8) | 24 (54.5) | 3 (20.0) | **0.034** |
| **Duration of symptoms (days)** |  |  |  |  |
| Median (IQR) | 6 (3, 9) | 6 (3, 10) | 5 (3, 8) | 0.488 |
| **Duration between 1st positive COVID-19 test**<br>**and Mucormycosis diagnosis** |  |  |  | 0.751 |
| < 7 days | 2 (3.4) | 2 (4.5) | 0 (0.0) |  |
| 7-14 days | 20 (33.9) | 14 (31.8) | 6 (40.0) |  |
| 15-28 days | 28 (47.5) | 20 (45.5) | 8 (53.3) |  |
| > 28 days | 9 (15.3) | 8 (18.2) | 1 (6.7) |  |
| **Median (IQR)** | **17 (12, 22)** | **17 (13, 23)** | **17 (10, 21)** | **0.676** |

IQR- Interquartile range

**Table 4:**
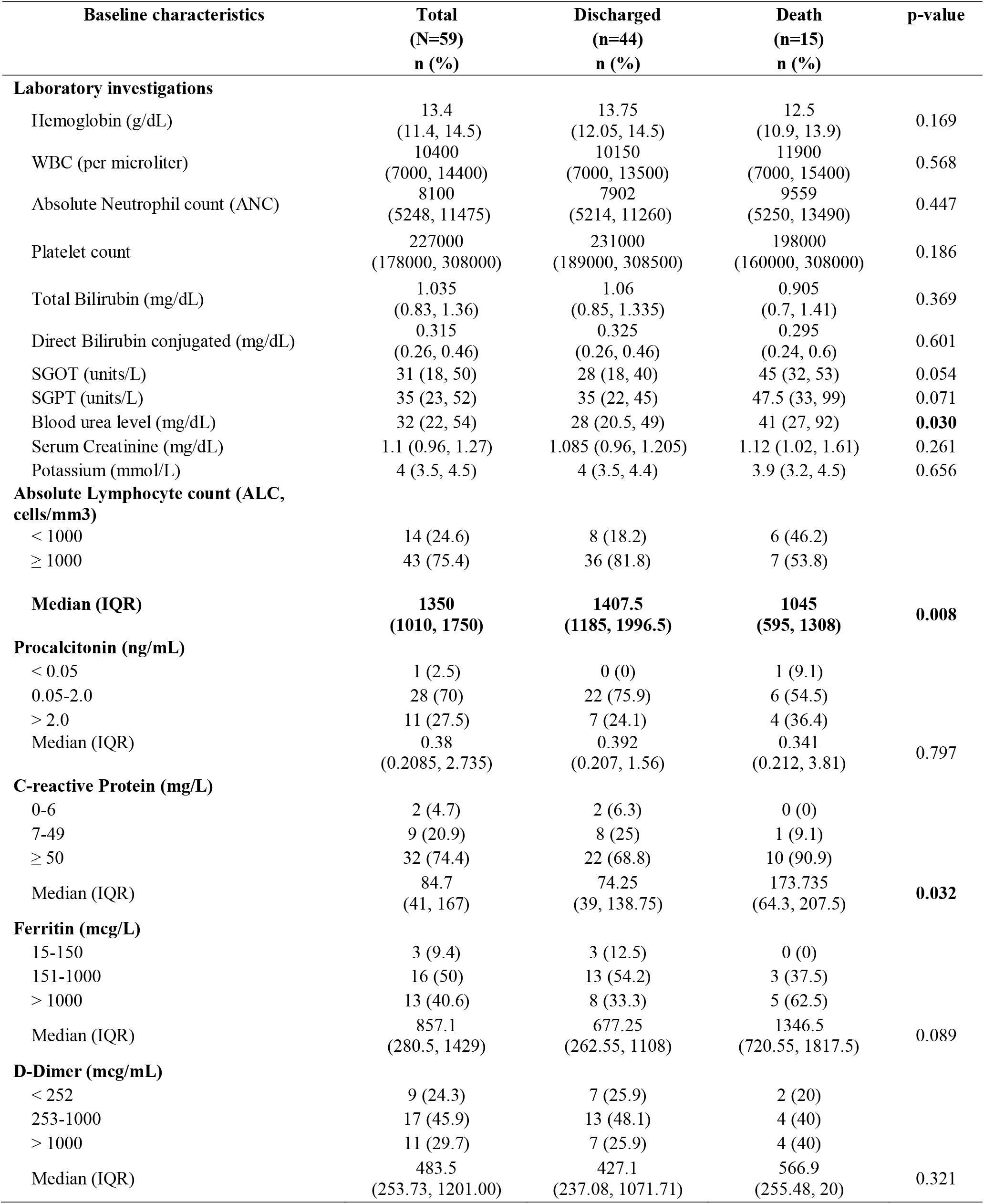
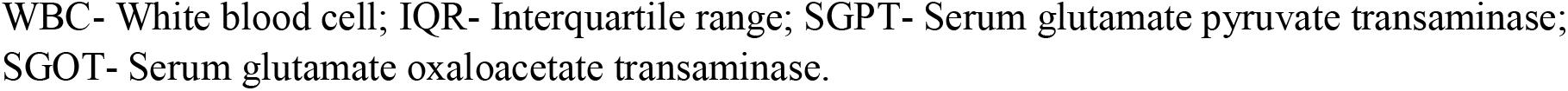
Baseline investigations during admission for Mucormycosis.

Aggressive surgical debridement of involved site and combination antifungal therapy was standard of care for all ROCM patients in our cohort. Empirical combination anti-fungal therapy was started after radiologic evidence of invasive sinusitis, pending confirmatory diagnosis by histopathology or culture. Various treatment modalities used in our cohort are mentioned in Table 5. Intravenous (IV) formulations of Amphotericin B (Liposomal, Lipid-complex, Lipid emulsion or Deoxycholate) were the mainstay of antifungal therapy and prescribed to 91.5% (54/59) patients. In view of shortage of Liposomal Amphotericin B in India during the second COVID-19 wave, patients ended up getting a combination of Amphotericin B formulations as per availability. Liposomal Amphotericin B was prescribed to 89.8 % (53/59) patients for a median duration of 15 (IQR: 11, 21) days. Amphotericin B de-oxycholate or Conventional Amphotericin B was prescribed to 45.8% (27/59) patients for a median duration of 4 (IQR: 3, 9) days. The median duration of Amphotericin B exposure to patients in our cohort was 21 (IQR: 14, 27) days. All patients were also prescribed concomitant oral triazole therapy (Posaconazole (58/59, 98.3%) or Isavuconazole (5/59, 8.5%)) during admission and after discharge till clinician was satisfied about local control of disease. Median duration for oral triazole therapy was 60 days. FESS with orbital decompression was performed on 94.9 % patients (56/59). Surgical exenteration of the eye with debridement of orbital cavity was performed in 13 (22 %) patients. Partial maxillectomy was performed in 9 (15.3 %) patients while Neurosurgery for frontal lobe brain abscess drainage was performed in 5 (8.5 %) patients. Eight patients (13.6 %) required invasive mechanical ventilation while 18 (30.5 %) required ICU admission for Mucormycosis.

**Table 5:**
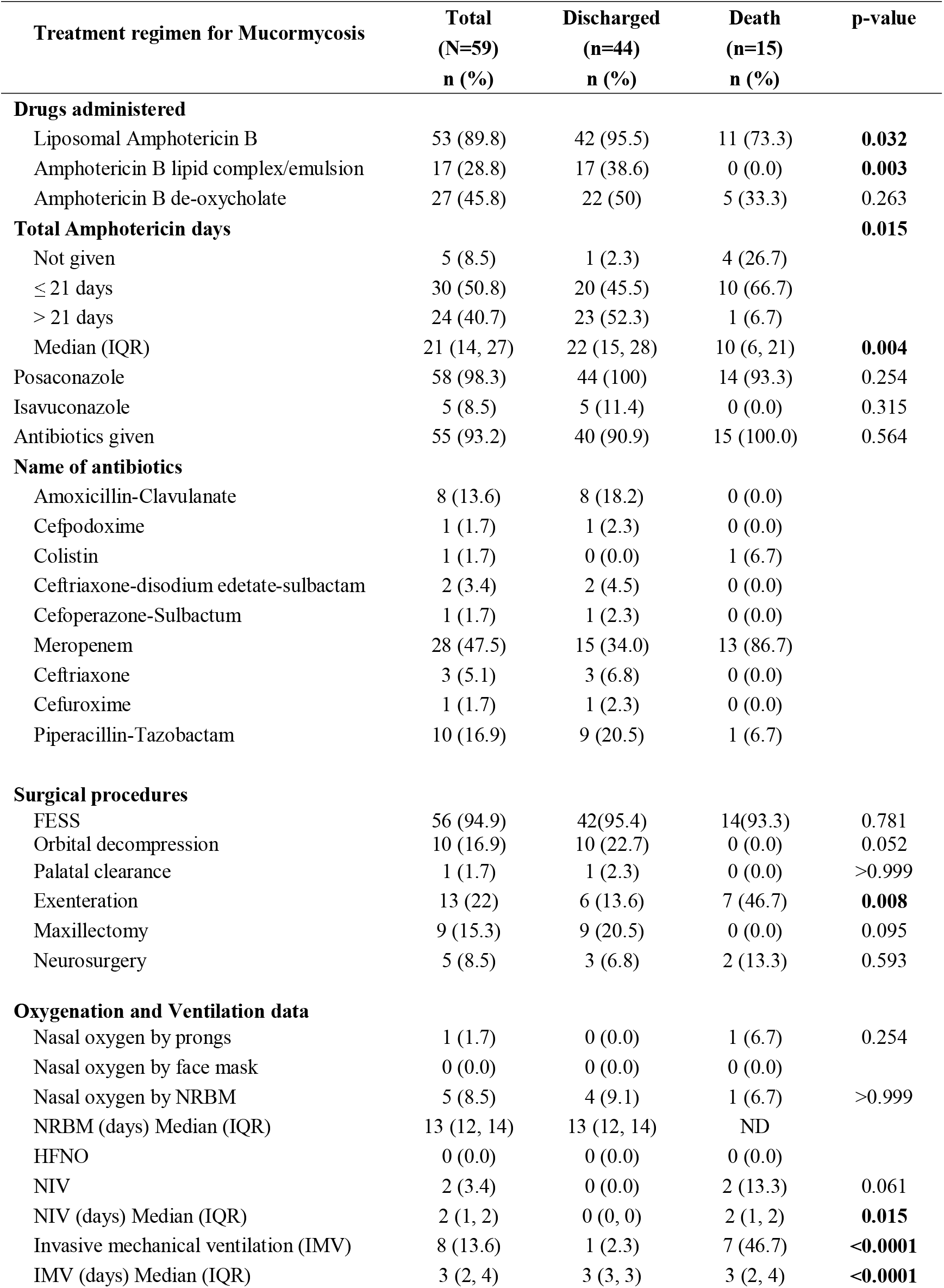
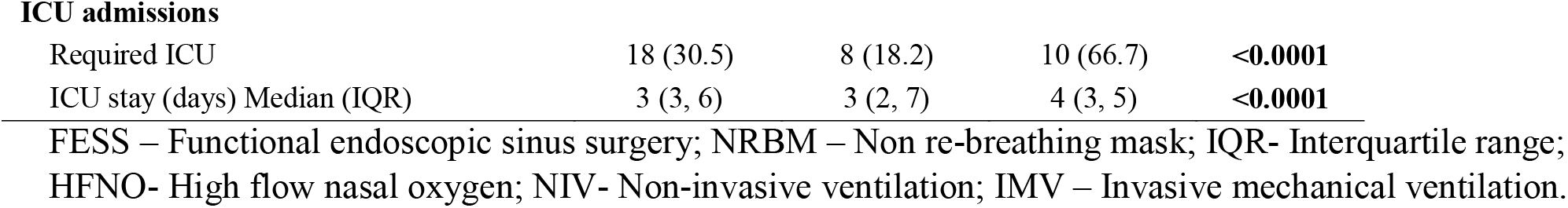
Treatment regimens used in Mucormycosis.

| Treatment regimen for Mucormycosis | Total<br>(N=59)<br>n (%) | Discharged<br>(n=44)<br>n (%) | Death<br>(n=15)<br>n (%) | p-value |
| --- | --- | --- | --- | --- |
| **Drugs administered** |  |  |  |  |
| Liposomal Amphotericin B | 53 (89.8) | 42 (95.5) | 11 (73.3) | **0.032** |
| Amphotericin B lipid complex/emulsion | 17 (28.8) | 17 (38.6) | 0 (0.0) | **0.003** |
| Amphotericin B de-oxycholate | 27 (45.8) | 22 (50) | 5 (33.3) | 0.263 |
| **Total Amphotericin days** |  |  |  | **0.015** |
| Not given | 5 (8.5) | 1 (2.3) | 4 (26.7) |  |
| ≤ 21 days | 30 (50.8) | 20 (45.5) | 10 (66.7) |  |
| > 21 days | 24 (40.7) | 23 (52.3) | 1 (6.7) |  |
| Median (IQR) | 21 (14, 27) | 22 (15, 28) | 10 (6, 21) | **0.004** |
| Posaconazole | 58 (98.3) | 44 (100) | 14 (93.3) | 0.254 |
| Isavuconazole | 5 (8.5) | 5 (11.4) | 0 (0.0) | 0.315 |
| Antibiotics given | 55 (93.2) | 40 (90.9) | 15 (100.0) | 0.564 |
| **Name of antibiotics** |  |  |  |  |
| Amoxicillin-Clavulanate | 8 (13.6) | 8 (18.2) | 0 (0.0) |  |
| Cefpodoxime | 1 (1.7) | 1 (2.3) | 0 (0.0) |  |
| Colistin | 1 (1.7) | 0 (0.0) | 1 (6.7) |  |
| Ceftriaxone-disodium edetate-sulbactam | 2 (3.4) | 2 (4.5) | 0 (0.0) |  |
| Cefoperazone-Sulbactam | 1 (1.7) | 1 (2.3) | 0 (0.0) |  |
| Meropenem | 28 (47.5) | 15 (34.0) | 13 (86.7) |  |
| Ceftriaxone | 3 (5.1) | 3 (6.8) | 0 (0.0) |  |
| Cefuroxime | 1 (1.7) | 1 (2.3) | 0 (0.0) |  |
| Piperacillin-Tazobactam | 10 (16.9) | 9 (20.5) | 1 (6.7) |  |
| **Surgical procedures** |  |  |  |  |
| FESS | 56 (94.9) | 42(95.4) | 14(93.3) | 0.781 |
| Orbital decompression | 10 (16.9) | 10 (22.7) | 0 (0.0) | 0.052 |
| Palatal clearance | 1 (1.7) | 1 (2.3) | 0 (0.0) | >0.999 |
| Exenteration | 13 (22) | 6 (13.6) | 7 (46.7) | **0.008** |
| Maxillectomy | 9 (15.3) | 9 (20.5) | 0 (0.0) | 0.095 |
| Neurosurgery | 5 (8.5) | 3 (6.8) | 2 (13.3) | 0.593 |
| **Oxygenation and Ventilation data** |  |  |  |  |
| Nasal oxygen by prongs | 1 (1.7) | 0 (0.0) | 1 (6.7) | 0.254 |
| Nasal oxygen by face mask | 0 (0.0) | 0 (0.0) | 0 (0.0) |  |
| Nasal oxygen by NRBM | 5 (8.5) | 4 (9.1) | 1 (6.7) | >0.999 |
| NRBM (days) Median (IQR) | 13 (12, 14) | 13 (12, 14) | ND |  |
| HFNO | 0 (0.0) | 0 (0.0) | 0 (0.0) |  |
| NIV | 2 (3.4) | 0 (0.0) | 2 (13.3) | 0.061 |
| NIV (days) Median (IQR) | 2 (1, 2) | 0 (0, 0) | 2 (1, 2) | **0.015** |
| Invasive mechanical ventilation (IMV) | 8 (13.6) | 1 (2.3) | 7 (46.7) | **<0.0001** |
| IMV (days) Median (IQR) | 3 (2, 4) | 3 (3, 3) | 3 (2, 4) | **<0.0001** |
| **ICU admissions** |  |  |  |  |
| Required ICU | 18 (30.5) | 8 (18.2) | 10 (66.7) | **<0.0001** |
| ICU stay (days) Median (IQR) | 3 (3, 6) | 3 (2, 7) | 4 (3, 5) | **<0.0001** |

FESS – Functional endoscopic sinus surgery; NRBM – Non re-breathing mask; IQR- Interquartile range; HFNO- High flow nasal oxygen; NIV- Non-invasive ventilation; IMV – Invasive mechanical ventilation.

Complications during antifungal therapy and hospital admission are mentioned in Table 6. Fever with chills (78 %), Thrombophlebitis (59.3 %), Hypokalemia (88.1 %), Anemia (57.6 %) and Nephrotoxicity (50.8 %) were the most common complications observed during hospital admission. Central nervous system (CNS) complications (stroke/cerebritis/brain abscess) were seen in 25.4 % patients. There were 14 deaths (23.7 %) in our cohort during first hospital admission for Mucormycosis. On Relative risk analysis, CT severity score during COVID-19 admission _≥_ 18 (p = 0.017), presence of symptoms like eye swelling (p = 0.002), loss of vision (p< 0.0001), ptosis (p = 0.005) and limb weakness (p = 0.001), presence of diabetic ketoacidosis (p = 0.011), cerebral involvement by Mucor (p = 0.0004) and development of CNS complications like stroke (p < 0.0001) during antifungal therapy were associated with increased risk of death (Table 7, Figure 1). Duration of Amphotericin B treatment of more than 21 days was associated with decreased risk of death (p = 0.002) on Relative risk analysis (Table 7). All patients who were discharged from hospital were followed up for a minimum period of 6 weeks to look for late complications. Three patients needed re-admission, 48, 54 and 37 days after first discharge in view of re-appearance of symptoms and/or imaging evidence of progression of disease to the CNS. All 3 patients needed repeat surgical debridement and additional Amphotericin B treatment. They were successfully discharged the second time. Two patients also needed re-admission for osteomyelitis of maxilla and alveolar arch which needed partial maxillectomy and placement of dental prosthesis. One patient who had undergone exentration of eye was readmitted 90 days later for insertion of orbital prosthesis. One patient who had been treated for cerebral mucormycosis (bilateral complete vision loss and multiple cerebral infarcts) with 42 days of high dose Liposomal Amphotericin B and Posaconazole was readmitted with aspiration pneumonia 12 days later and died. Cause of death was respiratory failure due to aspiration pneumonia and acute respiratory distress syndrome (ARDS, Total deaths in cohort - 15, 25.4%).

**Table 6:** Complications in Mucormycosis patients during antifungal therapy.

| Complications in<br>Mucormycosis | Total<br>(N=59)<br>n (%) | Discharged<br>(n=44)<br>n (%) | Death<br>(n=15)<br>n (%) | p-value |
| --- | --- | --- | --- | --- |
| **Complications** |  |  |  |  |
| Acute kidney injury | 30 (50.8) | 22 (50) | 8 (53.3) | 0.824 |
| Hypokalemia | 52 (88.1) | 40 (90.9) | 12 (80.0) | 0.355 |
| Hepatitis | 20 (33.9) | 12 (27.3) | 8 (53.3) | 0.066 |
| Fever | 46 (78.0) | 35 (79.5) | 11 (73.3) | 0.721 |
| Chills | 45 (76.3) | 34 (77.3) | 11 (73.3) | 0.757 |
| Loose motions | 14 (23.7) | 11 (25.0) | 3 (20.0) | >0.999 |
| Vomiting | 21 (35.6) | 15 (34.1) | 6 (40.0) | 0.680 |
| Loss of appetite | 46 (78.0) | 35 (79.5) | 11 (73.3) | 0.721 |
| Abdominal pain | 9 (15.3) | 5 (11.4) | 4 (26.7) | 0.213 |
| Abdominal Distension | 2 (3.4) | 1 (2.3) | 1 (6.7) | 0.447 |
| Thrombophlebitis | 35 (59.3) | 29 (65.9) | 6 (40.0) | 0.078 |
| Stomatitis | 20 (33.9) | 15 (34.1) | 5 (33.3) | 0.957 |
| Pedal edema | 14 (23.7) | 9 (20.5) | 5 (33.3) | 0.311 |
| Anemia | 34 (57.6) | 22 (50.0) | 12 (80.0) | 0.068 |
| Thrombocytopenia | 14 (23.7) | 6 (13.6) | 8 (53.3) | **0.002** |
| Osteomyelitis | 11 (18.6) | 10 (22.7) | 1 (6.7) | 0.259 |
| Stroke/ Cerebritis | 15 (25.4) | 2 (4.5) | 13 (86.7) | **<0.0001** |

**Table 7:**
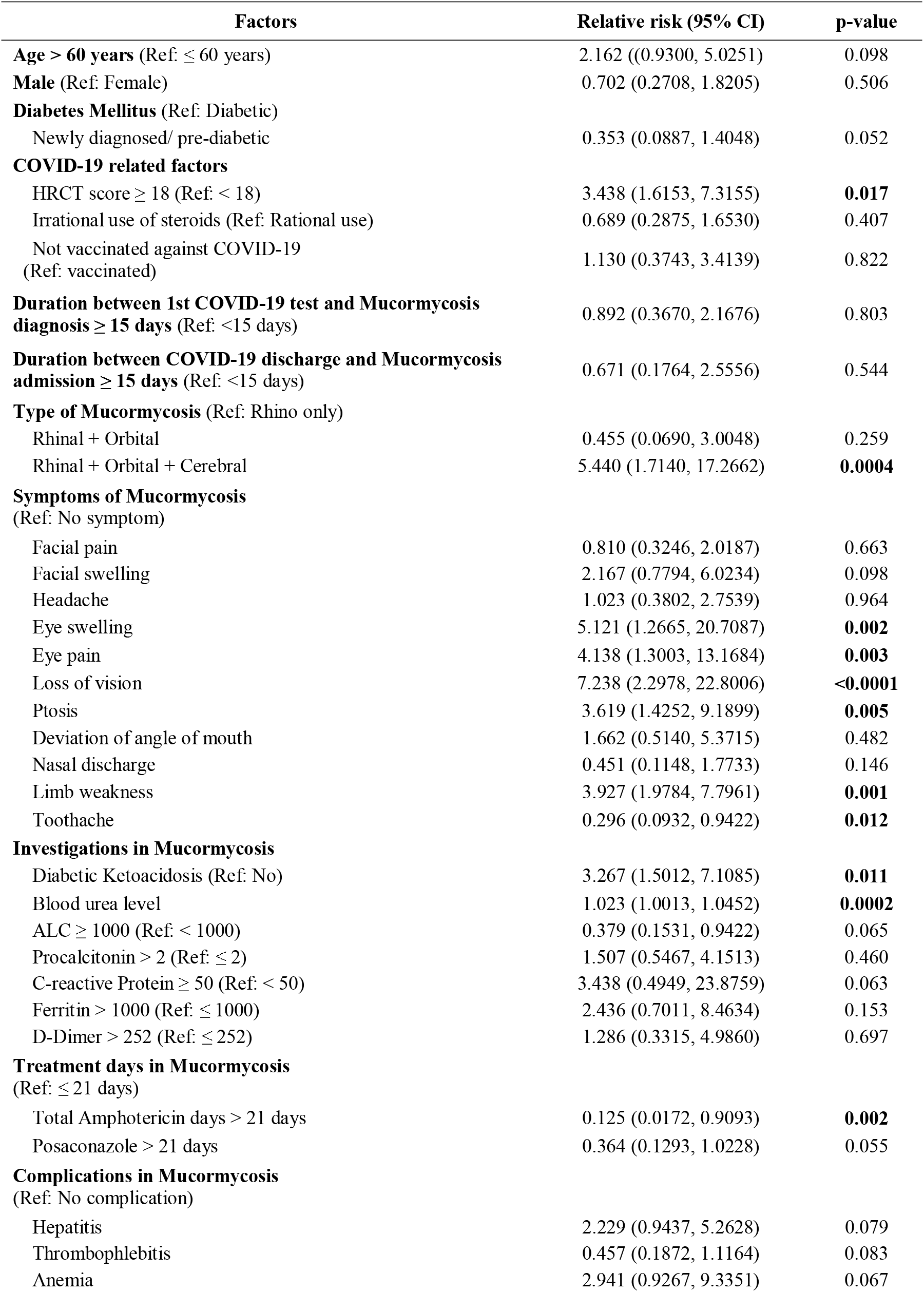
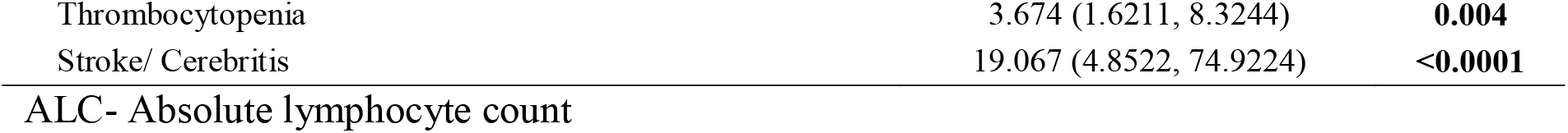
Relative risk of death in Mucormycosis patients.

**Figure 1:**
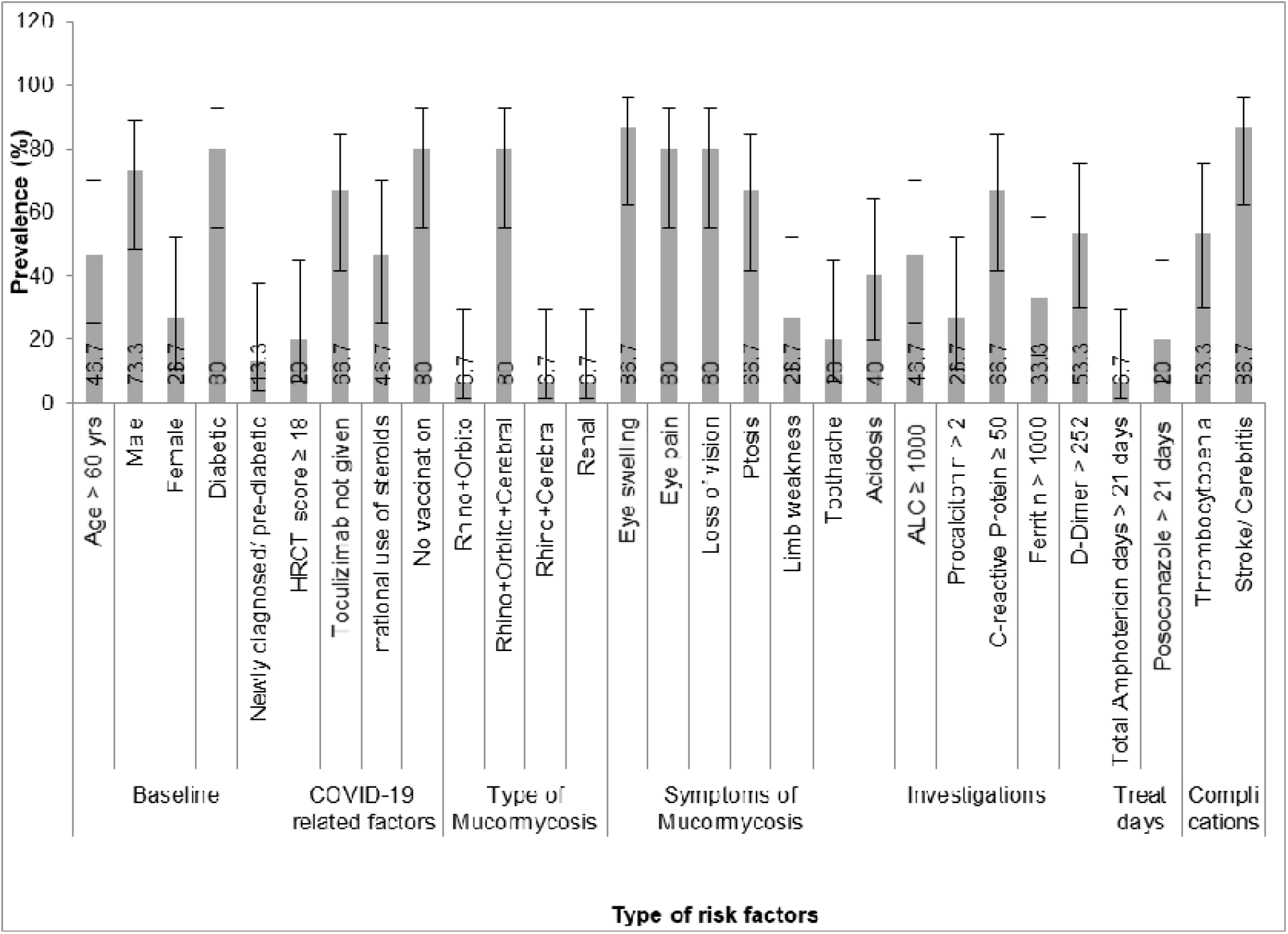
Prevalence of risk factors in Mucormycosis patients who died

## Discussion

We conducted a single-centre, retrospective cohort study of 59 patients with CAM who were admitted at NHRC in the last 15 months. 74.6% patients could be successfully discharged after control of disease with combined surgical and medical approach. Mortality rate in our cohort at 6 weeks follow-up was 25.4 %.

Majority of the patients in our study were middle-aged (median age: 52□years), of which nearly two-thirds were male. It has been hypothesized that the effect of estrogen might be protective in systemic fungal infection, which could have led to lower incidence in females ^[68]^. Case series on CAM by Sharma et al ^[46]^, described diabetes as a risk factor in 90% cases of which 52% had uncontrolled disease. A systematic review of 101 cases of CAM by Singh et al ^[58]^ also noted that more than 80% cases had either pre-existing or new onset hyperglycaemia as a risk factor. Our study has demonstrated similar findings in that 90 % of included patients were diabetic with almost 30% being newly diagnosed with DM at the time of admission for COVID-Of the 56 patients who had HbA1c value available, 50 had uncontrolled DM (HbA1c > 7%). Median HbA1c in our cohort was 9.3% while it was 9.6% in the study by Sen et al ^[55]^. Tight control of blood sugar level among patients with Diabetes Mellitus/ COVID-19 co-infection could help in reducing incidence of ROCM.

Both the RECOVERY trial ^[17]^ and WHO COVID-19 ^[69]^ guidelines clearly recommend against usage of Corticosteroids in COVID-19 patients not requiring oxygen (absence of hypoxia). Despite that, indiscriminate use of steroids in mild COVID-19 continues in India. The underlying reasons include the sudden surge of cases during second COVID wave leading to panic among general practitioners, inadequate time for triaging patients in busy outpatient clinics, non evidence-based clinical practice, availability of over the counter steroids, shortage of hospital beds and inadequate monitoring of the patients taking steroids. The improper use of corticosteroids has been identified as an independent risk factor for CAM by the MucoCovi network ^[7]^. In their retrospective analysis of 187 Indian CAM patients, they found that 78% (150/187) had received steroid therapy but only 33% patients had received steroids at appropriate dosages. In 33 % patients (50/150), steroids were not indicated while among 30% (45/150) patients steroids were indicated but were prescribed in inappropriately high dose ^[7]^. In comparison, in our study, 56 % patients were prescribed steroids for non-hypoxemic, mild to moderate COVID illness (irrational steroid therapy) while in 8.5 % patients steroids were indicated but were prescribed in inappropriately high dose. Thus there is an urgent need to stop prescription of steroids in non-hypoxemic COVID-19 patients and to limit the dose and duration of steroids in hypoxic patients.

In our cohort, majority of the patients were treated with a combination of surgical debridement (94%) and intravenous Amphotericin B therapy (91%). Mortality rate at 6 weeks follow-up was 25.4 % which is lower than what is reported by Pal et al ^[57]^ and Patel et al (MucoCovi network - 6 week mortality rate of 38.3 % and 12 week mortality of 45.7 %) ^[7]^. Reasons for lower mortality in our cohort as compared to the MucoCovi network ^[7]^ could be relatively younger population (median age 52 years versus 56.9 years), lower prevalence of hypoxia during COVID-19 (44 % versus 56 %), lower pulmonary involvement due to Mucormycosis (0 % versus 8.6 %), increased use of combination antifungal therapy (91% versus 50%) and higher use of combined surgical and medical approach (91% versus 71%). However, presence of diabetic ketoacidosis (10.9 % versus 8.6 %) and prevalence of cerebral involvement due to Mucor (44 % versus 23 %) was higher in our cohort than the MucoCovi study ^[7]^. Both the factors, presence of diabetic ketoacidosis (p = 0.011) and cerebral involvement with Mucor (p = 0.0004) were associated with higher risk of mortality on Relative risk analysis. In patients with cerebral involvement due to Mucor, mortality rate was 50% which is 5 times higher than among patients without cerebral involvement. On the other hand, duration of Amphotericin B therapy of more than 21 days was associated with statistically significant reduction in mortality (p = 0.002). Development of ROCM while the patient is still under active treatment for moderate or severe COVID-19 may pose significant challenges in the management – specifically termination of corticosteroids, and surgery under general anesthesia. This was reflected in the CT severity index > 18 (depicting severe COVID pneumonia) being a significant risk factor for increased risk of death due to CAM (p = 0.017). Patients with ROCM also remain at risk for delayed complications and may need re-admission with further surgical and anti-fungal treatment to resolve them.

### Limitations

Our study has several limitations. First, this is not a randomized controlled trial, and therefore unmeasured confounding cannot be ruled out. Second, as for all retrospective studies, some individuals diagnosed with Mucormycosis may be unreported leading to measurement bias and underestimation of mortality due to CAM. Third, we collected data from a single centre in India unlike other multicenter cohort studies ^[7, 52,54,55,57]^. Fourth, an overwhelmed health care system, inadequate workforce and lack of exhaustive reporting due to surge of cases during second COVID wave could be responsible for underestimation of co-morbidities, presenting symptoms and complications amongst patients in our cohort. Fifth, inflammatory markers like Ferritin, CRP and D-dimer were not available for all patients in the cohort. Sixth, we did not look for environmental factors causing healthcare-associated mucormycosis like contaminated ventilation systems, air conditioners, and ongoing construction in our hospital. We did not estimate the burden of Mucormycetes spores in our hospital environment. We also didn’t investigate the link between risk factors like use of industrial oxygen during the COVID pandemic, contaminated nebulizer fluids or inline humidifier tubing used in ventilator circuits and contaminated oxygen delivery systems with increased incidence of CAM in our cohort ^[10]^. Seventh, other unexplored factors, including genetic predisposition were not identified.

Despite these limitations, this retrospective cohort study adds to the growing body of literature on epidemiology, management strategies, outcomes and long term complications due to COVID-19 associated Mucormycosis (CAM).

## Conclusions

COVID-19 associated Mucormycosis (CAM) is an uncommon, rapidly progressive, angio-invasive, opportunistic fungal infection which is fatal if left untreated. The most common form of CAM seen in our cohort was the Rhino-orbito-cerebral (ROCM) one. COVID-19 induced immune dysregulation, uncontrolled diabetes mellitus and indiscriminate use of steroids may be the risk factors associated with the sudden surge in Mucormycosis cases in India during the second wave. Clinicians should have a high index of suspicion for ROCM in patients recovering from COVID-19, especially among patients with new or previously diagnosed diabetes mellitus and clinical manifestations of headache, facial or orbital pain. Combination of surgical debridement and intravenous Amphotericin B therapy leads to clinical and radiologic improvement in majority of cases. Cerebral involvement by Mucor is associated with higher mortality.

## Data Availability

All data related to the manuscript is available at Figshare

https://figshare.com/s/b96305da2644d4b3de12

## Acknowledgements

Manisha Ghate MD, PhD (National AIDS Research Institute (NARI), Pune, India) - edited the manuscript.

## Abbreviations

SARS-CoV-2: Severe acute respiratory syndrome coronavirus 2
COVID-19: coronavirus disease 2019
WHO: World health organization
CAM: COVID-19 associated Mucormycosis
CRP: C-reactive protein
LDH: lactate dehydrogenase
IL-6: Interleukin - 6
IFN-γ: Interferon Gamma
DM: diabetes mellitus
TCZ: Tocilizumab
CRS: cytokine release syndrome
DKA: diabetic ketoacidosis
ROCM: Rhino-orbito-cerebral mucormycosis
NHRC: Noble hospital and Research Centre
HRCT: High resolution computerized tomography scan
SpO_2_: oxygen saturation
PaO_2_/FiO_2;_: ratio of arterial partial pressure of oxygen to fraction of inspired oxygen
FESS: Functional endoscopic paranasal sinus surgery
LAm B: Liposomal Amphotericin B
RT-PCR: reverse transcription PCR
ICU: Intensive care unit
CNS: Central nervous system
IRB: institutional review board
IQR: interquartile range
ANC: absolute neutrophil count
ALC: absolute lymphocyte count
IV: Intravenous
ARDS: acute respiratory distress syndrome

